# Effectiveness of a New Regional Network on STEMI Care in an Under-developed Area of Southwest China

**DOI:** 10.1101/2023.02.03.23285433

**Authors:** Li Mei Zhang, Alan Frederick Geater, Heng Luo, Yuan Zhang Wang, Shao Chang Wen

## Abstract

**Background:** ST-elevation myocardial infarction (STEMI) is life-threatening and need time-critical care. A new prefecture-wide STEMI Network was implemented in Chuxiong, Yunnan, China, the first reported STEMI network in underdeveloped area. How the Network impacted STEMI care in the prefecture has not previously been evaluated. The study aims to estimate the efficacy of the STEMI Network.

**Methods:** A longitudinal study including 5-years STEMI patients covering the pre-network, creation and post-network phase was conducted to assess the changes of STEMI care. Outcomes including timely presentation, reperfusion therapy, timely reperfusion, heart failure, inpatient mortality, length of hospital stay, in-hospital charge, and various intervals of ischemic time (total ischemic time, patient delay, system delay, diagnosis time, reperfusion delay, Z to N time, Z to W time) were compared among the three Network phases.

**Results:** A total of 1436 STEMI patients (380 in pre-network, 375 in creation and 681 in post-network phases) were included. Significantly increasing proportions of timely presentation (71.3%, 73.3%, 81.4%) and reperfusion (58.2%, 59.2%, 65.3%) were found over the 3 phases. Compared with pre-network patients, post-network patients had shorter medians of patient delay (193 vs. 215 minutes), total ischemic time (348 vs. 380 minutes) and system delay (152 vs. 174 minutes). A significant decreasing trend of heart failure was observed (11.1%, 8.8%, 7.0%) across network phases. After conditioning, post-network patients were more likely than pre-network patients to have timely presentation (OR=1.70 [1.26, 2.30]), receive reperfusion therapy (OR=1.33 [1.02, 1.73]), and have shorter patient delay (HR=1.23 [1.08,1.40]) and total ischemic time (HR=1.22 [1.03,1.44]), but were less likely to receive timely reperfusion (OR=0.55 [0.36, 0.84]), timely PCI (OR=0.55 [0.34, 0.86), and shorter Z to W time (HR=0.78 [0.65, 0.94).

**Conclusion:** Improvements of STEMI reperfusion care by the regional Network were evident in this under-developed area; however, timely reperfusion care still needs to be enhanced.

## Introduction

In contrast to decreasing trends in developed countries (1), ischemic heart disease including ST-Elevation Myocardial Infarction (STEMI) is becoming a more common cause of death in the developing world such as India (2). Research from China (3) also reported that there was an increasing trend of mortality from ischemic heart disease in China between 2004 and 2010. A retrospective analysis including 13,815 STEMI patients in China (4) reported that, between 2001 and 2011, the estimated national rates of hospital admissions for STEMI per 100 000 people increased from 3.5 in 2001 to 7.9 in 2006, and 15.4 in 2011 (p<0.0001); Despite an increase in the use of primary PCI, the proportion of patients who did not receive reperfusion did not significantly change (45.3% in 2001 *vs* 44.8% in 2011, p=0.69), and the adjusted in-hospital mortality did not significantly change between 2001 and 2011 (odds ratio 0.82, p=0.07).

STEMI networks (5) were introduced to developed countries in the beginning of 21^st^ century. Several studies showed that the network could improve accessibility to invasive diagnosis and increased reperfusion treatment rates (6), resulting in reduced mortality among patients (7). However, implementation of a STEMI network system is still a huge challenge in middle- and low-income countries such as China. It is reported that the first regional collaborative chest pain rescue network in China was established by Guangzhou General Hospital in 2011. Two rescue networks in Guangzhou General Hospital and Shanghai Chest Hospital were accredited by the American-based Society of Cardiovascular Patient Care. The integrated regional network for STEMI care (IRN-STEMI) (8) was implemented in Xiamen prefecture of Fujian province in March 2013.

Notably, Guangzhou, Shanghai and Xiamen are modern metropolitan areas with a high economic level, and all are in the eastern coastal areas of China. There is no published reports about STEMI network in underdeveloped areas of China or in other developing or under-developed countries.

### Study setting

Chuxiong prefecture is an economically underdeveloped area in Yunnan province, Southwest China. The total area of the prefecture is 28,438 square kilometres, more than 90% of the area is mountainous. According to the 2018 government report of Chuxiong prefecture (9), the resident population of Chuxiong prefecture was 2,654,798. The gross domestic product (GDP) of Chuxiong prefecture in the year 2018 was 102 billion Yuan (9), while Beijing (Capital city of China) GDP was 3 032 billion Yuan, Kunming (Capital city of Yunnan province) GDP was 500 billion Yuan (10).

There are 11 public hospitals offering “western medicine” in Chuxiong prefecture. The people’s hospital of Chuxiong prefecture is the only hospital which can provide around-the-clock primary PCI for STEMI patients in the prefecture, and it received the certification of the Chest Pain Centre (standard version) from the national headquarters in the year 2016, certifying that the prefecture hospital is capable of providing in-hospital “green channel care” which access bypassing emergency room (ER) and coronary care unit (CCU) in the PCI-centre for patients with acute chest pain. However, primary PCI is not available in other hospitals with county level.

On January 2017, by the local government policy, the Chuxiong prefecture started to implement a prefecture-wide STEMI network system. The new regional network in Chuxiong prefecture was implemented following the protocols of China’s Chest Pain Centre and with modification of standard STEMI network from European Society of Cardiology based on the real conditions of the local health system and population. As shown in Figure 1, the regional STEMI network is constructed with the People’s Hospital of Chuxiong prefecture as the core centre, local Emergency Systems as core elements, and 10 non-PCI hospitals in counties and other primary care units in town and/or villages as branch elements. The Network also includes public education and professional training. Regional information sharing and prehospital care were achieved through remote transmission of real-time “Electrocardiogram (ECG) cloud” transmission technology and the use of WeChat mobile application software.

**Figure 1.**
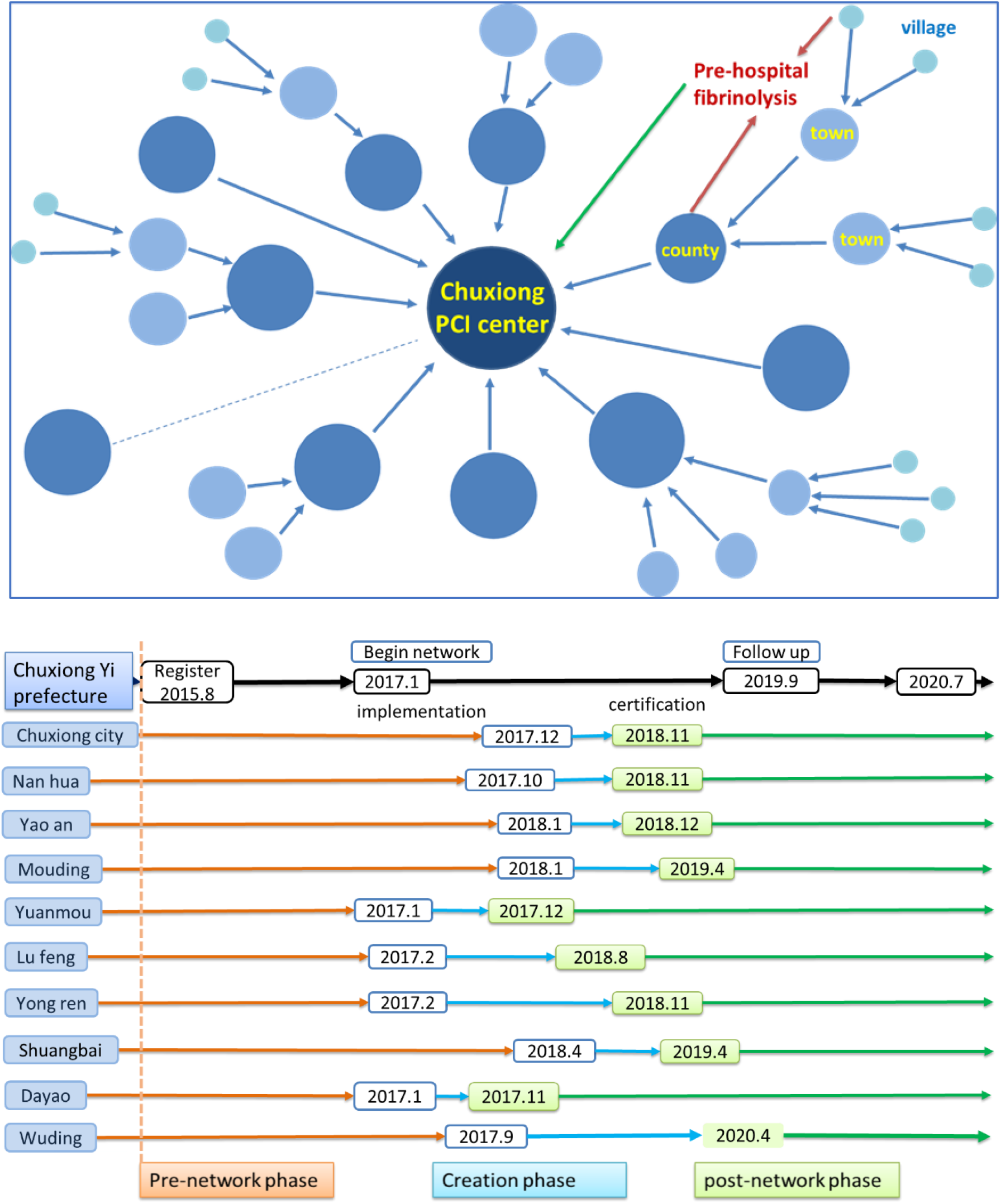
Conceptual structure and time frame of construction of the new regional STEMI network in Chuxiong, Yunnan, China

The implementation of a Chest Pain Centre (basic version) in a county hospital is a sign of entering a regional STEMI network. A requirement for certification is that the county hospital should cover all town hospitals in the county. Another requirement is that the hospital pass the on-site inspection, which is conducted by the headquarters of China Chest Pain Centre. It is notable that all the counties in Chuxiong prefecture received certification before April 2020. In other regions of China, only few of the local hospitals implemented chest pain centres (standard or basic version) in the administrative districts.

### Data Collection and Assessment Variables

After receiving the ethical approval from the Chest Pain Centre of Chuxiong prefecture, Yunnan, China, and also from the Human Research Ethic Committee in the Prince of Songkhla University, Thailand, the STEMI registry system database was searched to access STEMI index cases between August 2015 and July 2020, which includes 3 phases of the Network (pre-, creation phase, and post-). Then the STEMI cases were linked with medical records without name. Social determinants of health were collected including age (=< 50, 51-60, 61-70, >70 years), sex, ethnicity (Han majority, Yi or other minority), education (none, primary, secondary, higher), Occupation (farmer, others). Behavioural characteristics were collected, including insurance scheme, and smoking and drinking behaviour. Body mass index (BMI) was calculated based height and weight; BMI<18.0 is defined as underweight, 18-24.5 normal BMI, BMI > 24.5 overweight. Clinical characteristics were collected including symptoms (typical or atypical), onset period (day is defined as 8:am-6:00pm while night is defined as 6:00pm-8:am), mode of transport to hospital (by their own transportation or by ambulance), heart rate, respiratory rate, blood pressure, pulse oxygen, heart function (Killip class).

Regarding categorical outcome variables, timely presentation is defined as patients presenting with STEMI within 12 hours after onset of symptoms (the time from onset to receiving ECG diagnosis is less than or equal 12 hours). Reperfusion is defined as receiving prompt PCI (primary PCI or routine PCI) or emergency fibrinolysis. Timely fibrinolysis is defined as receiving fibrinolysis within 10 minutes after first diagnosis of ECG (only for patient who received fibrinolysis). Timely PCI is defined as receiving prompt PCI (primary PCI or routine PCI) within 90 minutes after diagnosis of ECG (only for patients who received prompt PCI). Timely reperfusion is defined as either timely fibrinolysis or timely PCI (for patient who received reperfusion). Major adverse cardiac event refers heart failure. Inpatient mortality was calculated for each phase of the Network.

Regarding continuous outcome variables, time from chest pain onset to first medical contact is defined as patient delay, time from first medical contact to first diagnosis by ECG is defined as diagnosis delay; for patient who received reperfusion, time from chest pain onset to receiving successful reperfusion is defined as total ischemic time, time from first medical contact to receiving successful reperfusion is defined as system delay, time from first diagnosis by ECG to receiving successful reperfusion (either emergency fibrinolysis or prompt PCI) is defined as reperfusion delay. The time between first diagnosis (time zero) and needle time for patients receiving fibrinolysis successfully is defined as Z to N time, while the time between first diagnosis (time zero) and wire crossing for patients receiving primary PCI is defined as Z to W time. Days and total charge in hospital also were collected.

### Statistical Analysis

All data analyses were performed using R and/or Stata.

Descriptive statistics were presented using means and standard deviations or medians and interquartile ranges for continuous variables where appropriate. Categorical variables were presented using frequencies and percentages. Comparison of categorical outcome variables among three phases of the Network was initially tested using the Chi-square test. Comparison of continuous outcome variables among three phases of the Network was initially tested using the Wilcoxon rank sum test where appropriate. Difference of ischemic time intervals (such as patient delay, system delay and total ischemic time) were also estimated using Kaplan-Meier analysis and Log-rank test. Competing risks analysis was employed to compare the cumulative incidence of reperfusion strategies among Network phases.

A Directed Acyclic Graph (DAG) was compiled depicting the hypothesized pathways influencing each outcome of interest (as shown in Figure I in supplementary file). Residence area, age, ethnicity and sex are evident as the minimal adjustment sets for estimating the total effect of Network on STEMI care. The minimal adjustment sets from DAG were included in all multivariate models. The total effects of the Network on STEMI care categorical outcomes were estimated using multivariate logistic regression models. The total effects of the Network on STEMI care continuous outcomes were estimated using Cox regression models.

## Results

A total of 1436 STEMI patients (380 during the pre-network phase, 375 during the creation phase and 681 during the post-network) were included.

### Demographic and Clinical Baseline Characteristics

Demographic and behavioral characters and clinical indices of all STEMI patients in each phase of network are shown in Table 1.

**Table 1.**
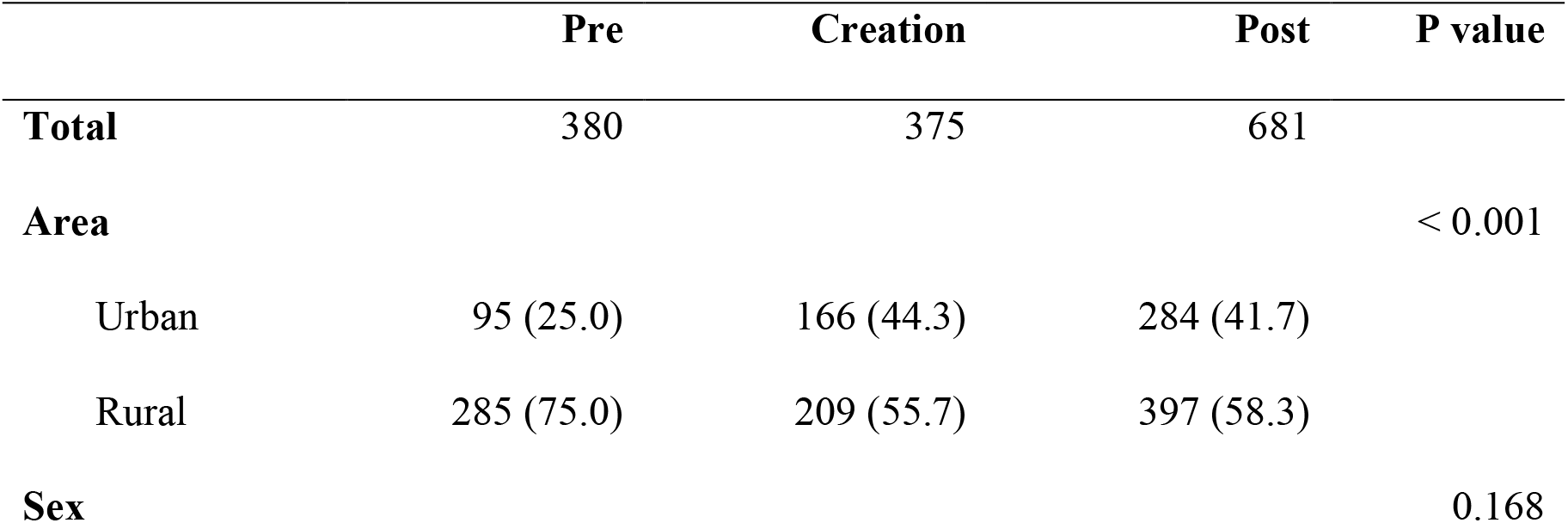
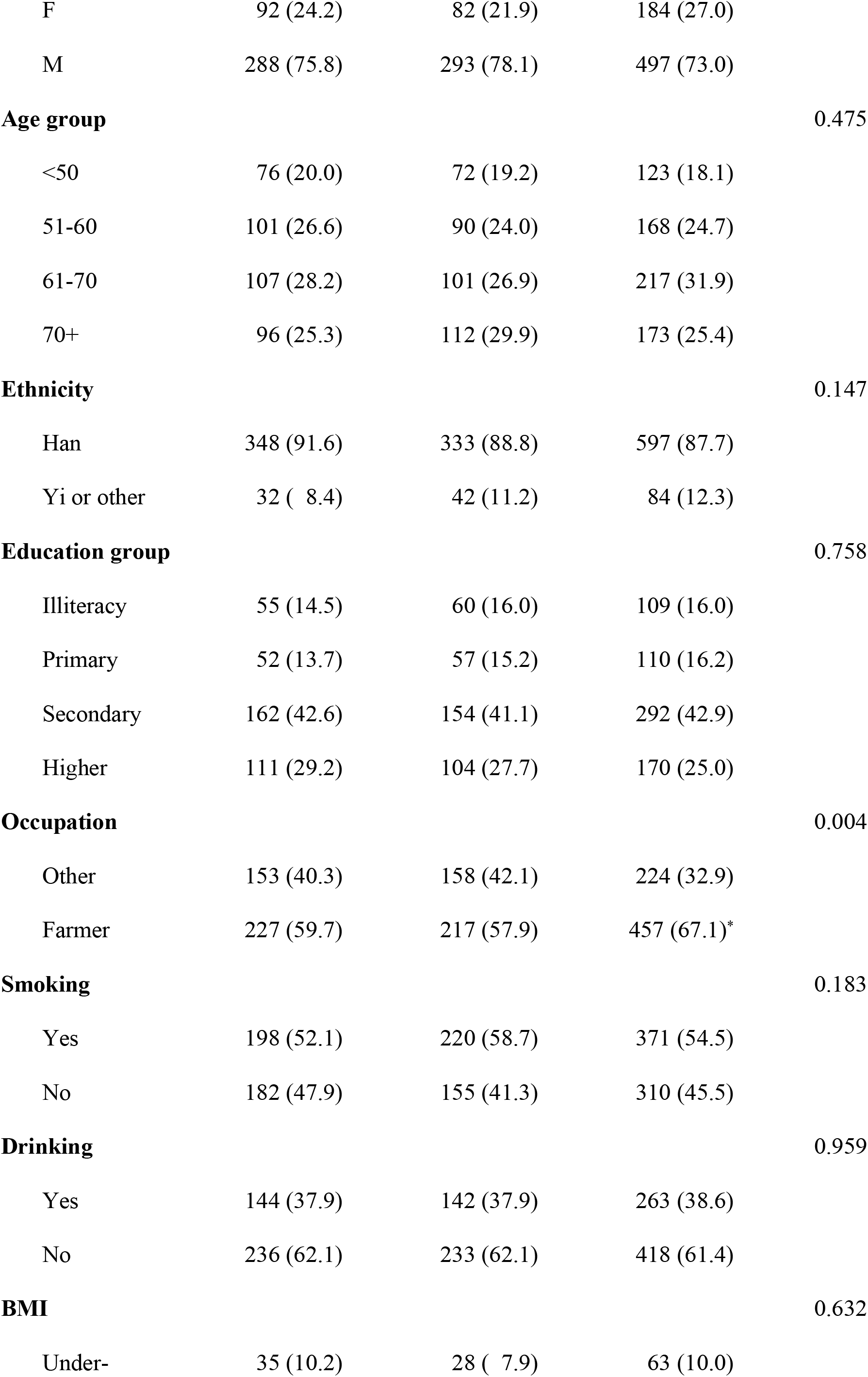
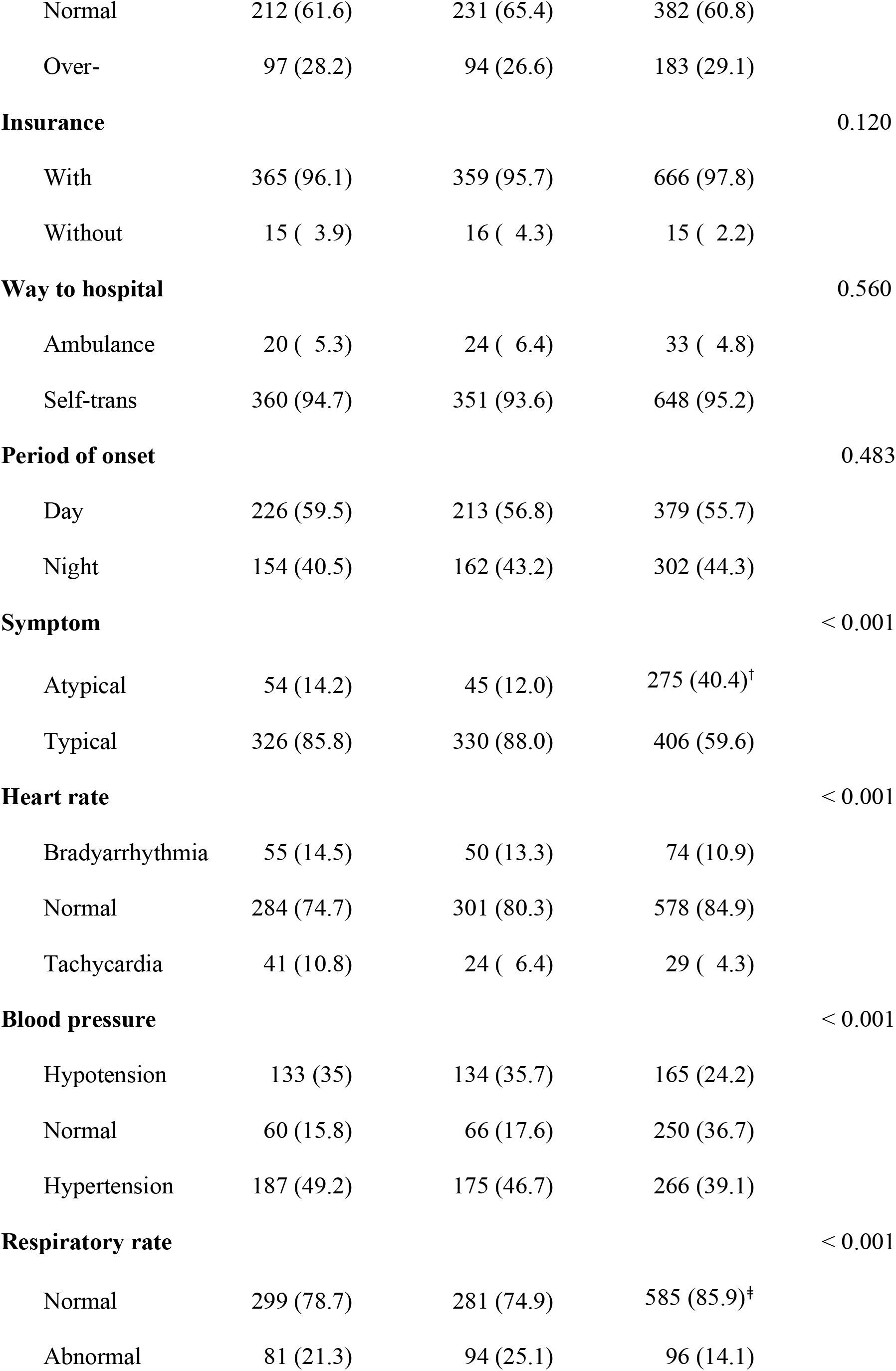
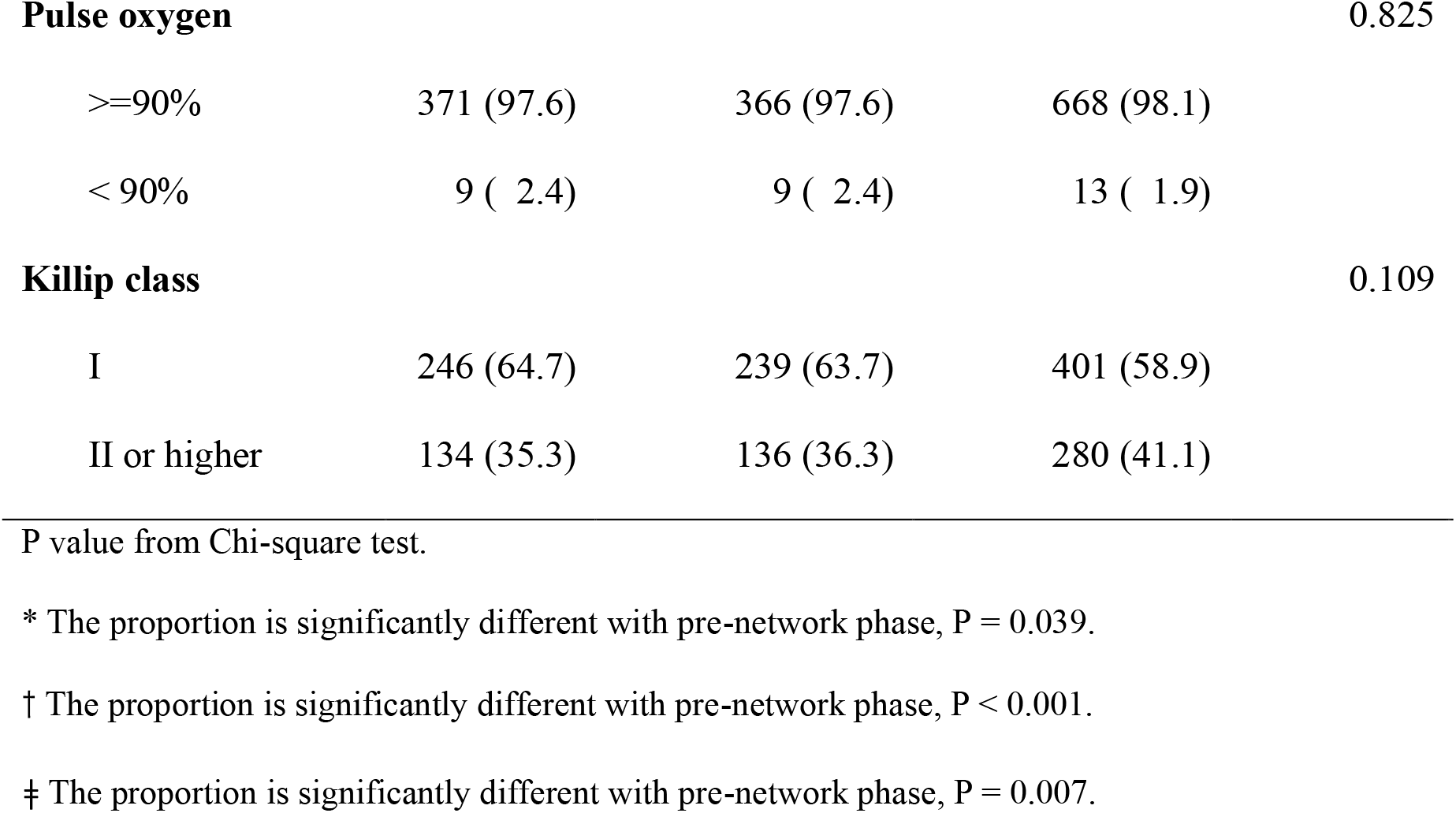
Baseline characteristics of 5-year STEMI patients

Among the patients who presented with STEMI in hospital, there were higher proportions of urban cases following the Network implementation (25.0%, 44.3%, 41.7%, respectively). Compared to patients during the pre-network phase, there were higher proportion of farmer (59.7% vs. 67.1%, P=0.039), with atypical symptom (14.2% vs. 40.4%, P<0.001), and with normal respiratory rate (78.7% vs. 85.9%, P=0.007) were observed in patients during the post-network phase. Increasing proportions of patients with normal heart rate (74.7%, 80.3%, 84.9%) and normal blood pressure (15.8%, 17.6%, 36.7%) were discovered from the pre-network phase to the post-network phase.

### STEMI Care Process and Proportions of Categorical Outcomes

The STEMI care process for all patients during each Network phase was shown in Figure 2. The proportions of STEMI care outcomes among three Network phases for all patients are shown with numbers in Table 2 and shown as histograms in Figure 3.

**Figure 2.**
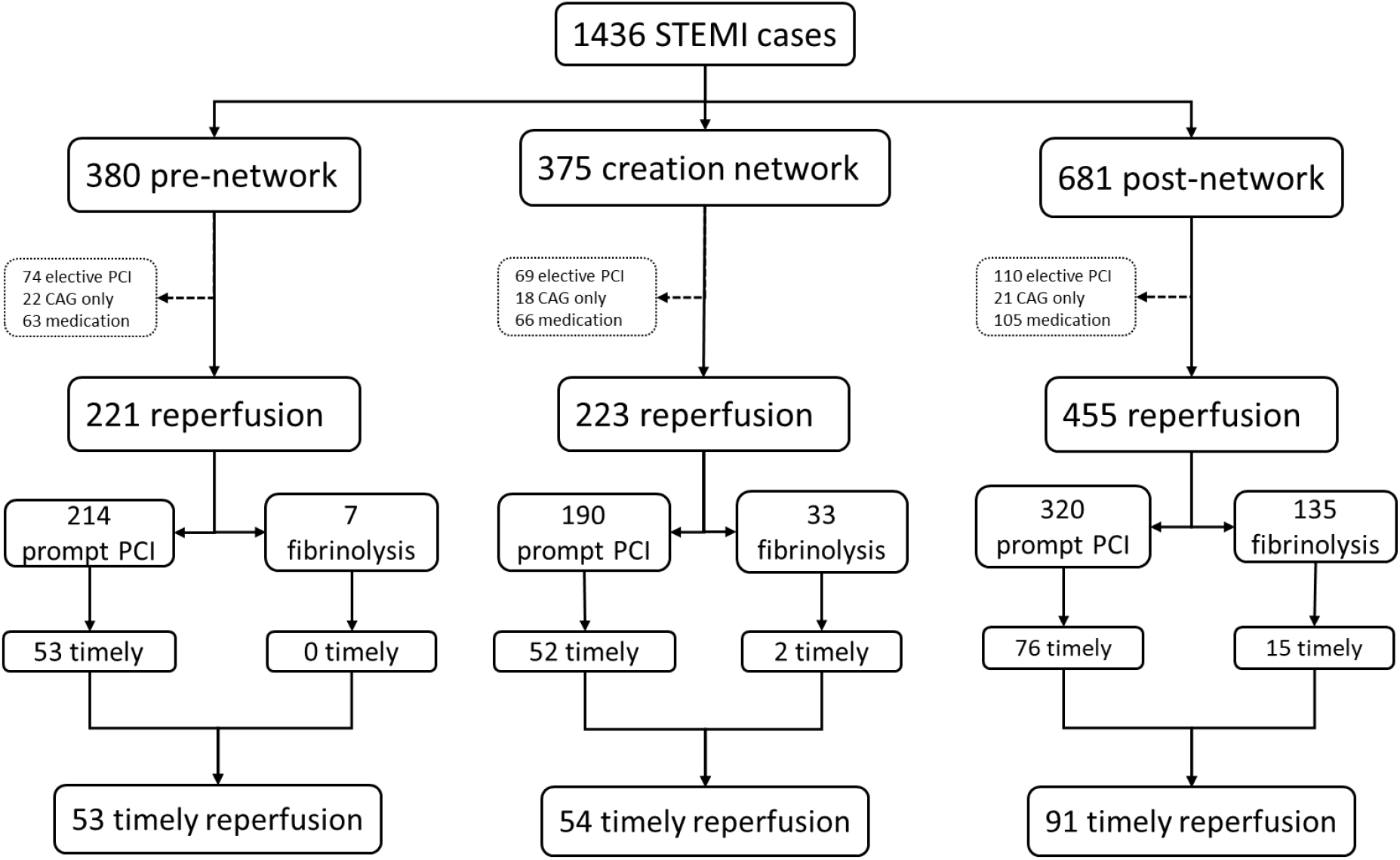
STEMI care process of all patients during Network phases

**Table 2.**
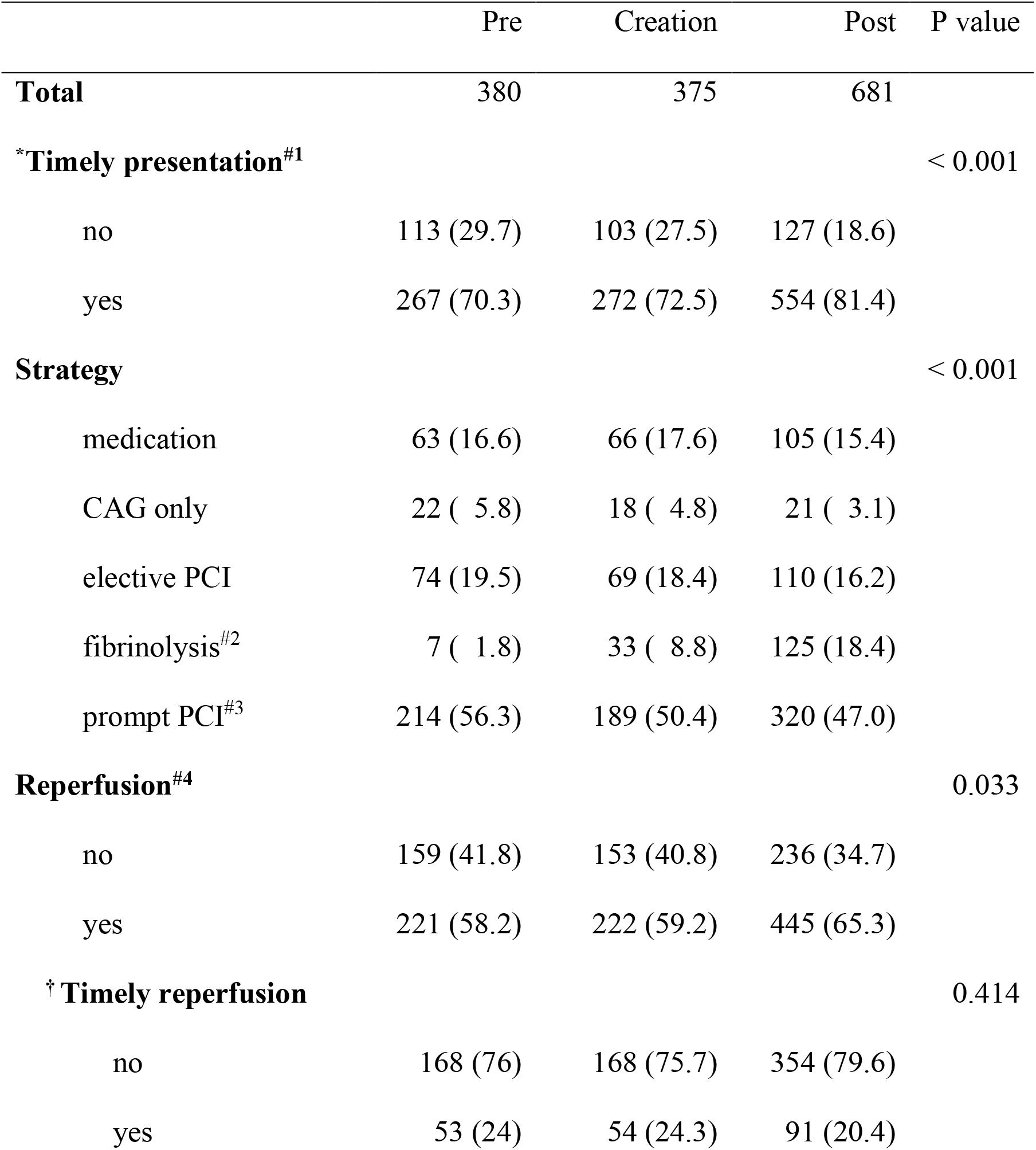
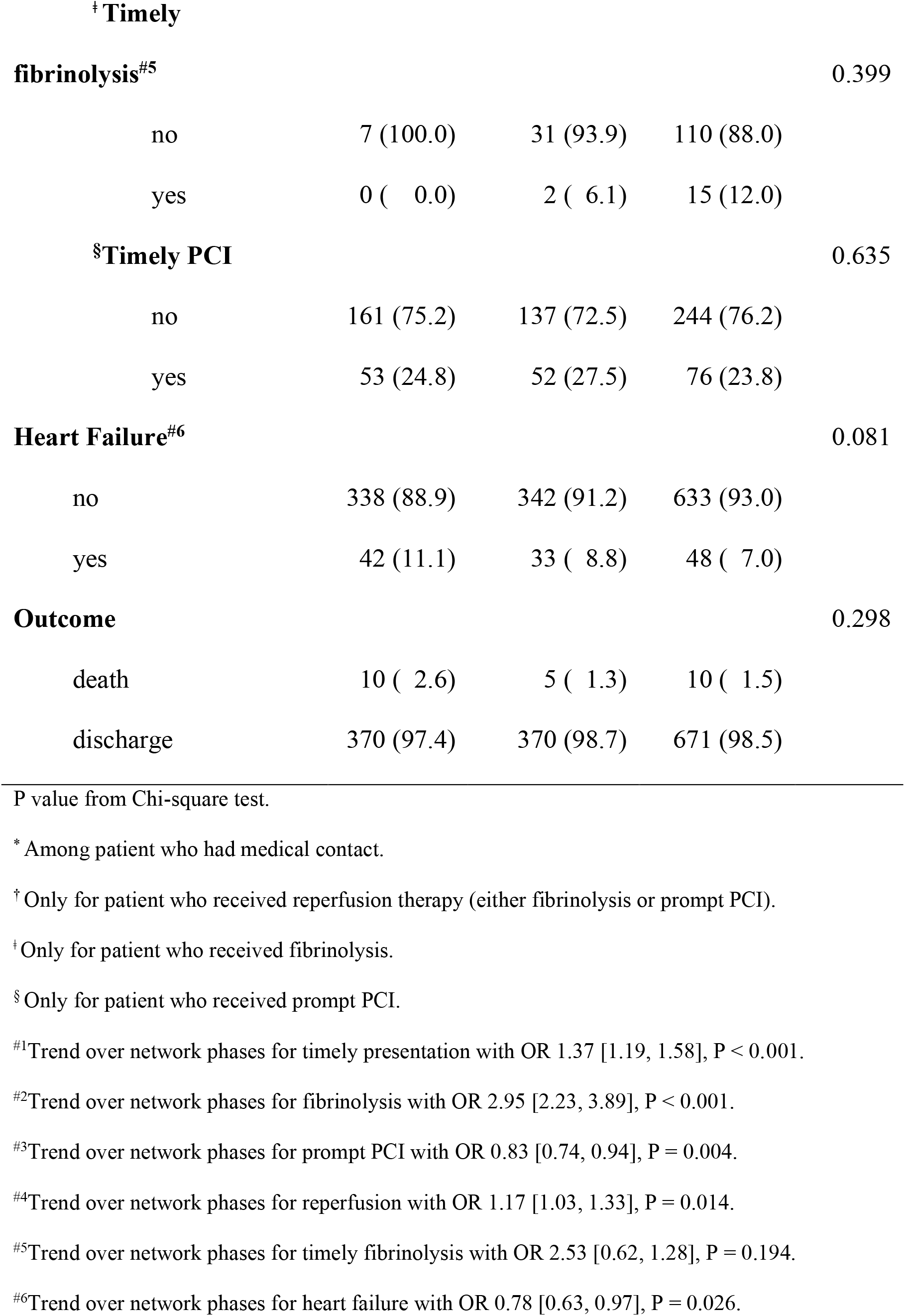
Univariate analysis of STEMI care outcomes for all patients

**Figure 3.**
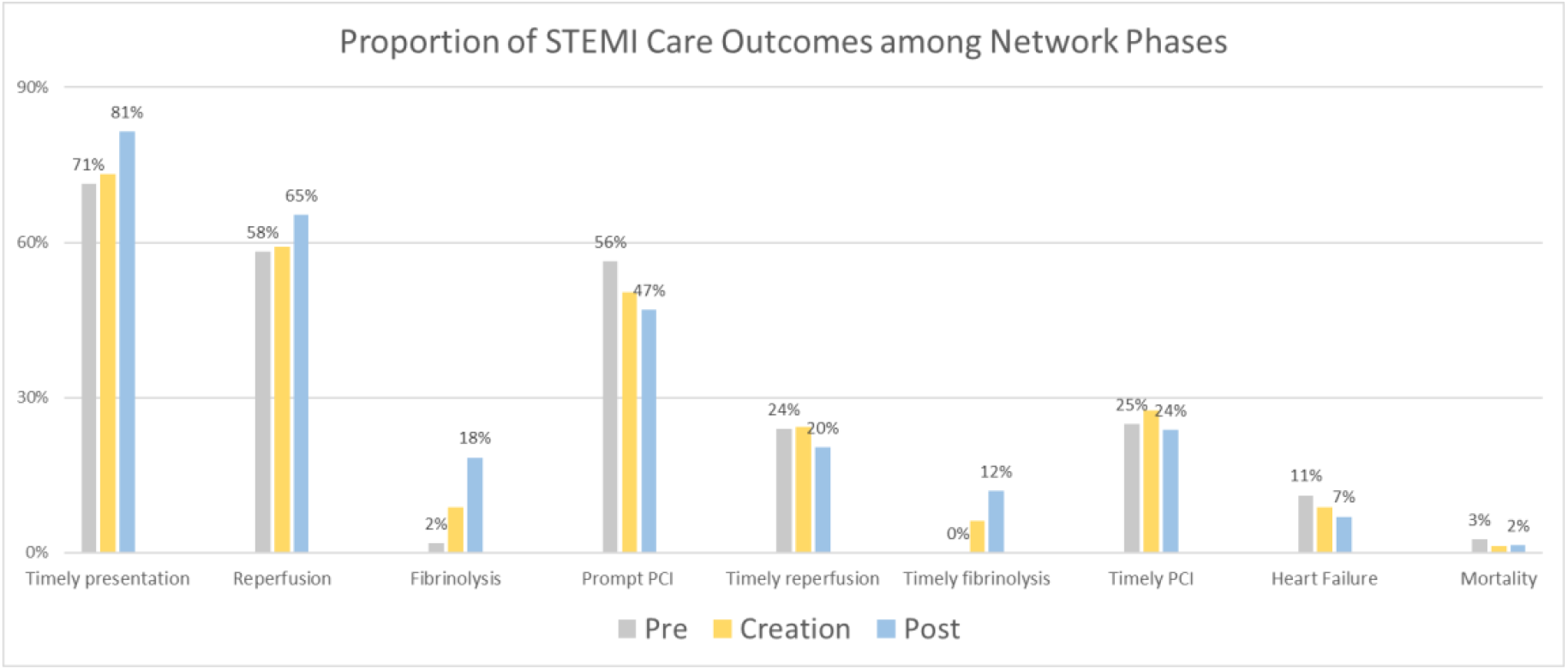
Proportions of STEMI care outcomes among Network phases

Among patients who presenting with STEMI in health care system, significantly increasing proportions of timely presentation were found from pre-network to post-network phase (70.3%, 72.5%, 81.4%) for all patients with an significant trend OR 1.37 [1.19, 1.58]. Among all patients, proportions underwent fibrinolysis were significantly increased from the pre-network to the post-network (1.8%, 8.8%, 18.4%; trend OR 2.95 [2.23, 3.89]) while proportions underwent prompt PCI were significantly decreasing from pre-network to post-network (56.3%, 50.4%, 47.0%; trend OR 0.83 [0.74, 0.94]). Totally, there were significantly increasing proportions of patients received reperfusion (either fibrinolysis or PCI) from the pre-network to the post-network phase (58.2%, 59.2%, 65.3%; OR 1.17 [1.03, 1.33]). Among those patients who received reperfusion (either fibrinolysis or PCI), the proportion of timely reperfusion was found to be stable across the network phases (24.0%, 24.3%, 20.4%).

Among those patients who received fibrinolysis, non-significant but increasing proportions of timely fibrinolysis were found from the pre-network phase to the post-network phase (0%, 6.1%, 12.0%; trend OR 2.53 [0.62, 1.28]). Among those patients who received PCI, the proportion of timely PCI were found to be stable across the network phases (24.8%, 27.5%, 23.8%).

Decreasing proportions of heart failure were found from pre-network to post-network (11.1%, 8.8%, 7.0%). Furthermore, there was a significant decreased trend across network phases for heart failure with OR 0.78 [0.63, 0.97]. There were non-significant differences of inpatient mortality for all patients among the three phases (2.6%, 1.3%, 1.5%).

### Odds Ratios of STEMI Care Categorical Outcomes by Network

Multivariate logistic models of STEMI care categorical outcomes are shown in Table 3.

**Table 3.**
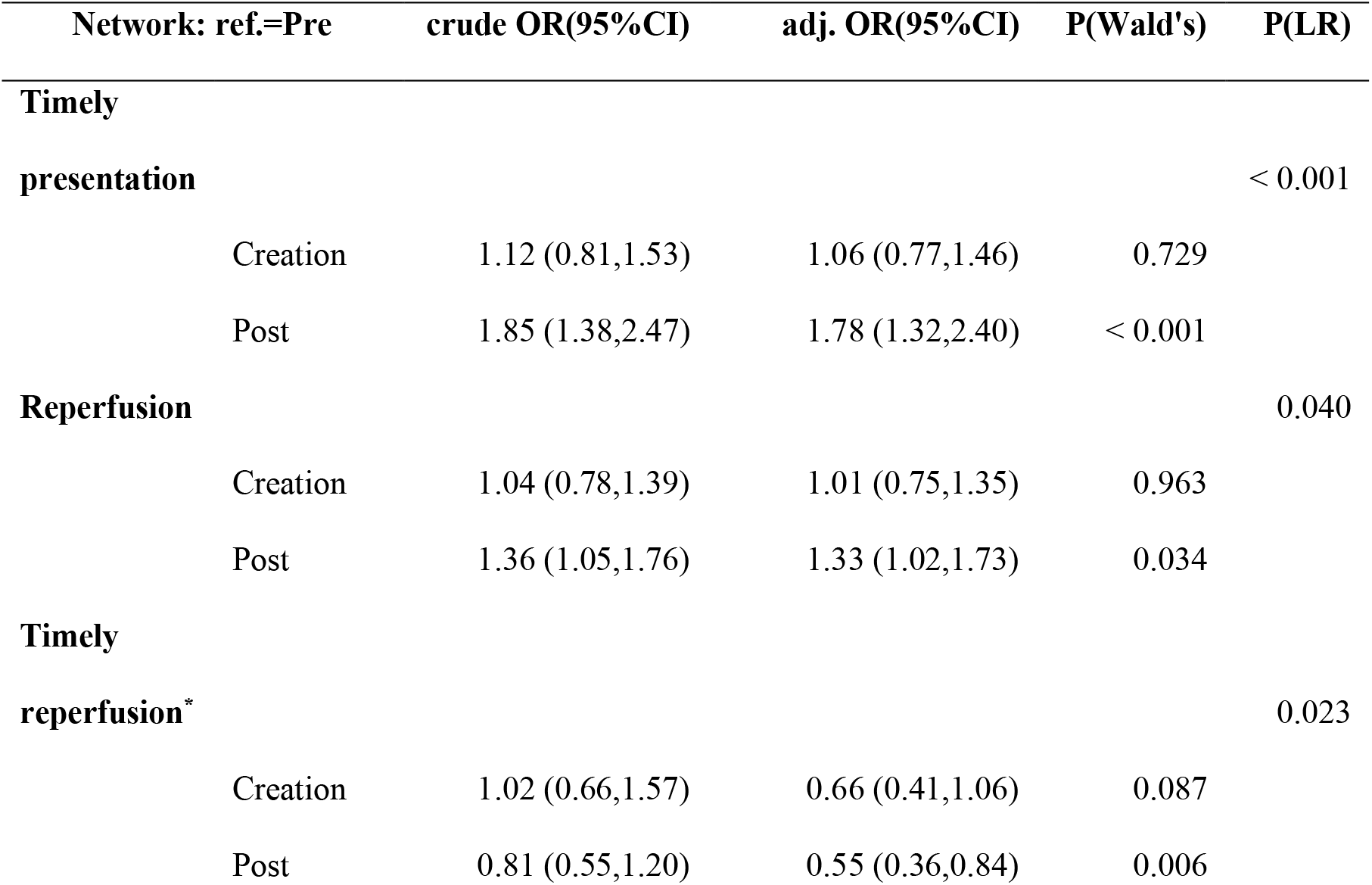
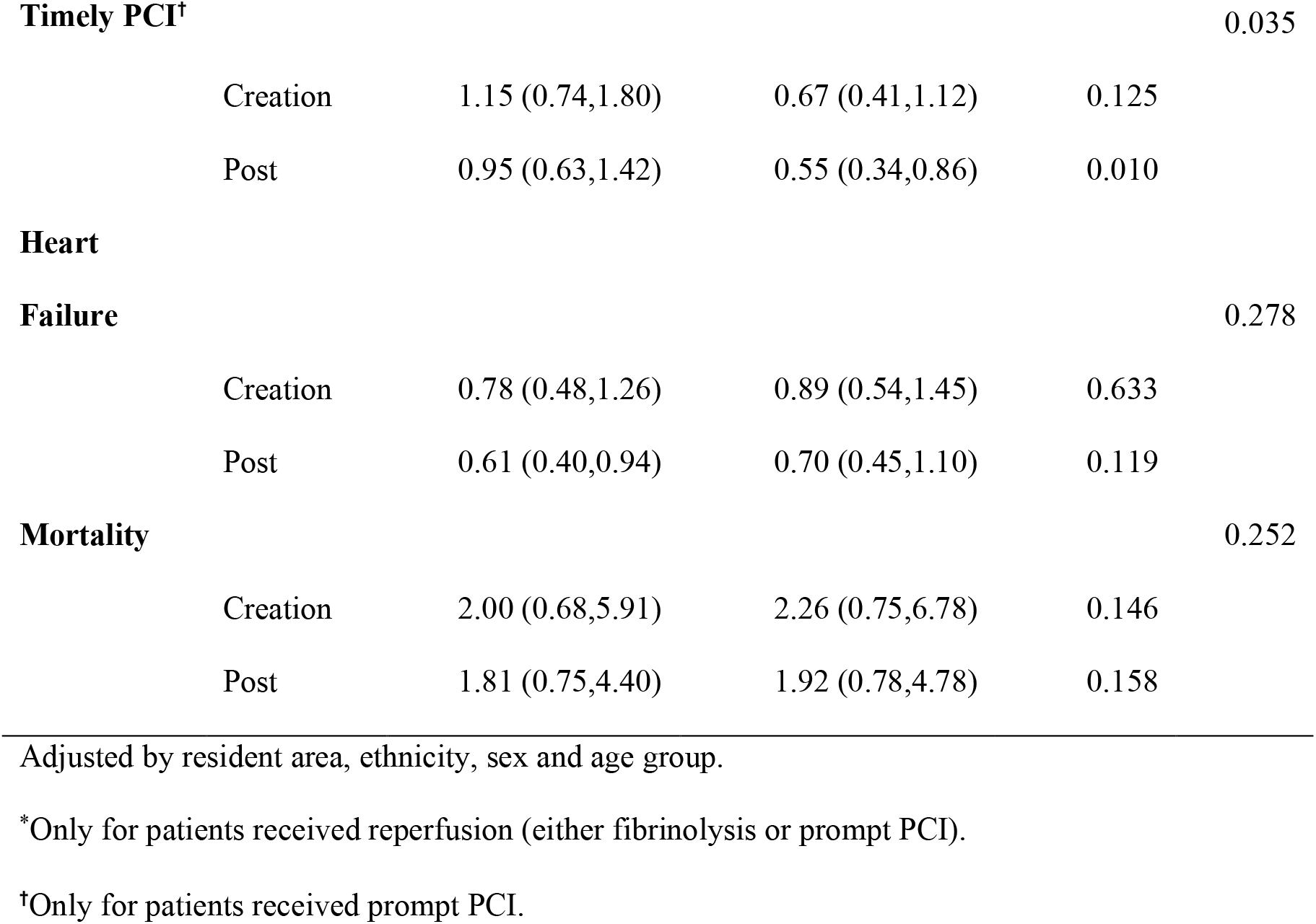
Logistic regression of STEMI care outcomes for all patients

After conditioning by resident area, ethnicity, sex and age group, patients during the post-network phase were significantly more likely to have timely presentation (OR=1.78 [1.32,2.40]) and receive reperfusion therapy (OR=1.33 [1.02, 1.73]) than patients during the pre-network.

However, among patients who received reperfusion (either fibrinolysis or prompt PCI), the group during the post-network phases were significantly less likely to receive timely reperfusion than the group during the pre-network phase (OR=0.55 [0.36, 0.84]). Among patients who received prompt PCI, the group during the post-network phases were significantly less likely to receive timely PCI than the group during the pre-network phase (OR=0.55 [0.34, 0.86]). For all patients, there were no significant difference of heart failure or inpatient mortality among the three phases of the network.

### Medians of STEMI Care Continuous Outcomes

Medians of all ischemic time intervals are summarized in Table 4. The median patient delay for patients during the post-network phase was significantly shorter than patients during the pre-network phase (192 minutes vs. 215 minutes, P<0.05). The median diagnosis time for patients during the post-network phase was significantly longer than patients during the pre-network phase (5 minutes vs. 4 minutes, P<0.05).

**Table 4.** Continuous outcomes of 5-year STEMI patients

| **Outcomes** | **Median (IQR) (minutes)** |  |  |  |
| --- | --- | --- | --- | --- |
|  | **Pre** | **Creation** | **Post** | **P** |
| Patient delay | 215.0 ( 82.5,855.8) | 225.0 ( 95.0,927.5) | 192.0 (70.0,487.0)* | 0.004 |
| Diagnosis time | 4.0 ( 1.0, 8.0) | 6.0 ( 4.0, 8.0) | 5.0 ( 4.0, 7.0)* | < 0.001 |
| Z to N time<sup>†</sup> | 24.0 ( 18.0,144.5) | 43.0 ( 16.0,109.0) | 38.0 ( 17.0, 86.0) | 0.866 |
| Z to W time <sup>‡</sup> | 157.5 ( 92.0,263.0) | 156.0 ( 86.0,268.0) | 180.5 ( 94.8, 285.5) | 0.300 |
| Reperfusion delay<sup>§</sup> | 155.0 ( 89.0,260.0) | 142.0 ( 73.0,246.5) | 133.5 ( 68.2,253.8)* | 0.052 |
| System delay<sup>§</sup> | 174.0 (103.0,290.0) | 152.0 ( 81.5,265.0) | 139.5 ( 74.2,268.0)* | 0.002 |
| Total ischemic time<sup>3</sup> | 380.0 (224.0,842.0) | 400.0 (235.5,664.0) | 347.5 (204.2,600.0)*,† | 0.021 |
| **Days in hospital** | 8.0 ( 6.0, 11.0) | 8.0 ( 6.0, 10.0) | 8.0 ( 6.0, 10.0) | 0.044 |
| **Charge (10,000 Yuan)** | 2.9 ( 2.1, 3.5) | 2.9 ( 1.9, 3.5) | 2.9 ( 2.4, 3.6) | 0.549 |

P value from Kruskal-Wallis test.
\* Significantly different with pre-network at the 0.05 level from pairwise Wilcox test.
<sup>†</sup> Only for patients received fibrinolysis.
<sup>‡</sup> Only for patients received prompt PCI.
<sup>§</sup> For patients received reperfusion (either fibrinolysis or prompt PCI).

For patients who received reperfusion (either fibrinolysis or prompt PCI), the median total ischemic time during the post-network phase was significantly shorter than during the pre-network phases (380 vs. 348 minutes); the median system delay showed a significant shortening trend from the pre-network phase to the post-network phase (174 minutes, 152 minutes and 140 minutes), and the median reperfusion delay during the post-network phase was significantly shorter than during the pre-network phase (155 vs. 134 minutes). For patients who received fibrinolysis, there were no significant difference of median Z to N time among the three phases of the network (24, 43, 38 minutes). For patients who received prompt PCI, there were no significant difference of median Z to W time among the three phases of the network (158, 156, 181 minutes). It is noticed that significantly narrower range of diagnosis time (5 [4, 7] vs. 4 [1, 8]) and days in hospital (8 [6, 10] vs. 8 [6, 11]) were observed during the post-network phase compared with the pre-network phase.

### Time-to-event curves for intervals of total ischemic time

The whole pictures of the performance of all patients’ STEMI care among the three network phases were compared using time-to-event curves in Figure 4.

**Figure 4.**
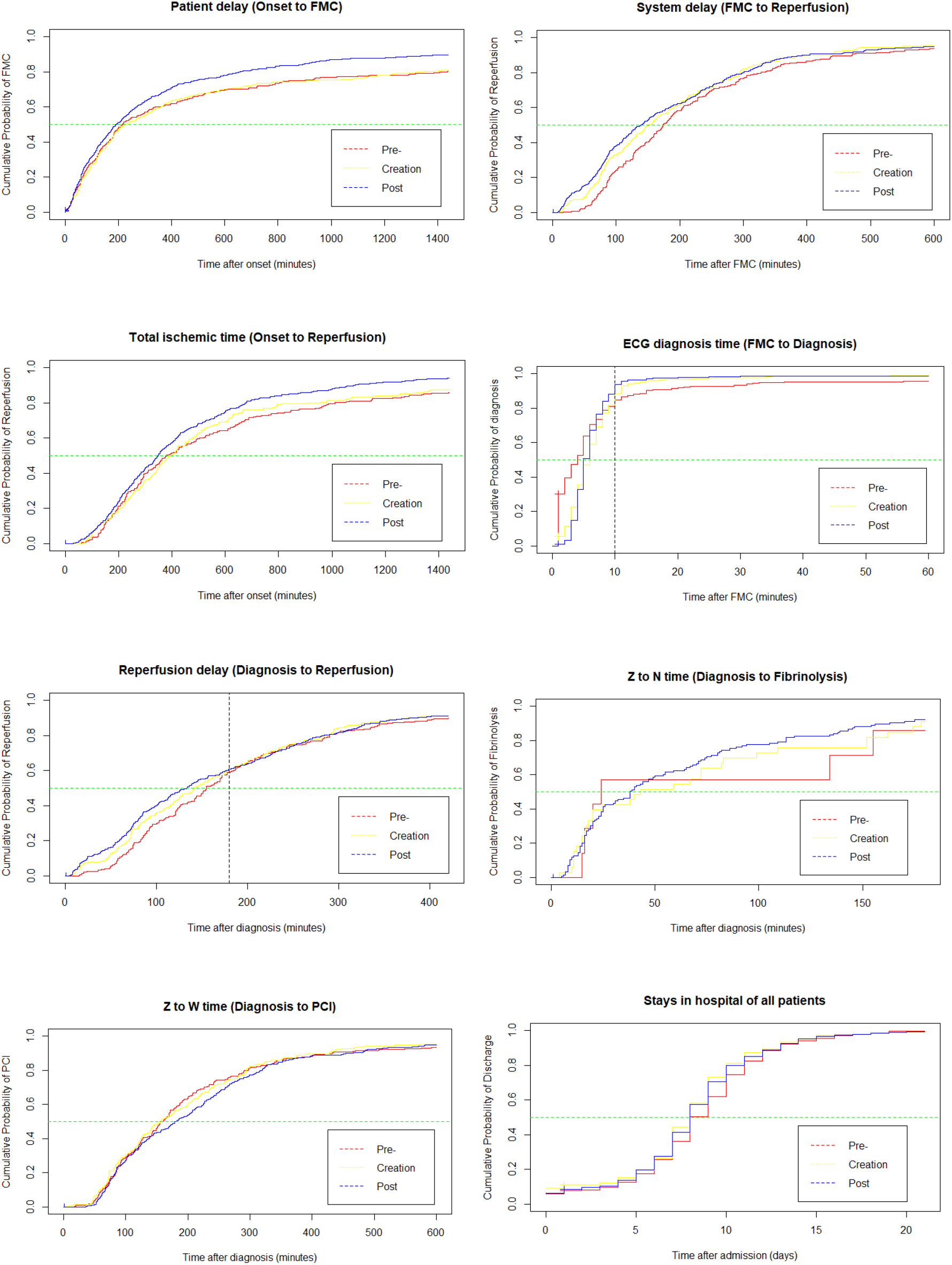
Time-to-Event Curves for Intervals of Ischemic Time The Y axis is the cumulative probability of reperfusion, X axis is the time inn minutes. Reperfusion delay, system delay and total ischemic time are only for patients received reperfusion (either fibrinolysis or PCI). Z to N time is only for patients who received fibrinolysis. Z to W time is only for patients who received prompt PCI.

For all patients, Significant shortening was found in patient delay in the post-network phase compared with the pre-network phase. At the same time point after onset, patients during the post-network phase had higher cumulative probability of contacting medical service and receiving reperfusion therapy than in the pre-network phase. For example, by 3 hours after onset, 44% patients had contacted medical service during the pre-network phase, while 49% patients had contacted during the post-network phase. The cumulative probability table of patient delay are shown in the Table I in Supplementary file.

It is noticed that, for all patients who had medical contact in health care system, there was no significant difference of median diagnosis time among the three phases of Network. However, for those patients did not receive first diagnosis by ECG within 10 minutes, compared with the pre-network patients, the post-work patients had a higher probability to receive early diagnosis after 10 minutes.

For patients who received reperfusion, significant shortening of system delay and total ischemic time were evident in patients during the post-network phase compared with patients during the pre-network phase. The cumulative probability table of system delay and total ischemic time are shown in the Table II and III in Supplementary file. There was no significant difference of reperfusion delay between pre-network patients and post-network patients. However, within 180 minutes after diagnosis (time zero), at a same timepoint, the post-network patients had higher probability to receive reperfusion than pre-network patients.

There was no significant difference for other outcomes including Z to N time, Z to W time, days in hospital and the total charge among the network phases.

### Hazard Ratios of STEMI Care Continuous Outcomes by Network

Multivariate Cox models were used to evaluate the Hazard Ratios (HR) of ischemic time intervals during the creation/post-network phases compared with pre-network phase. The crude and adjusted HRs for each continuous outcome variables are shown in Table 5.

**Table 5.**
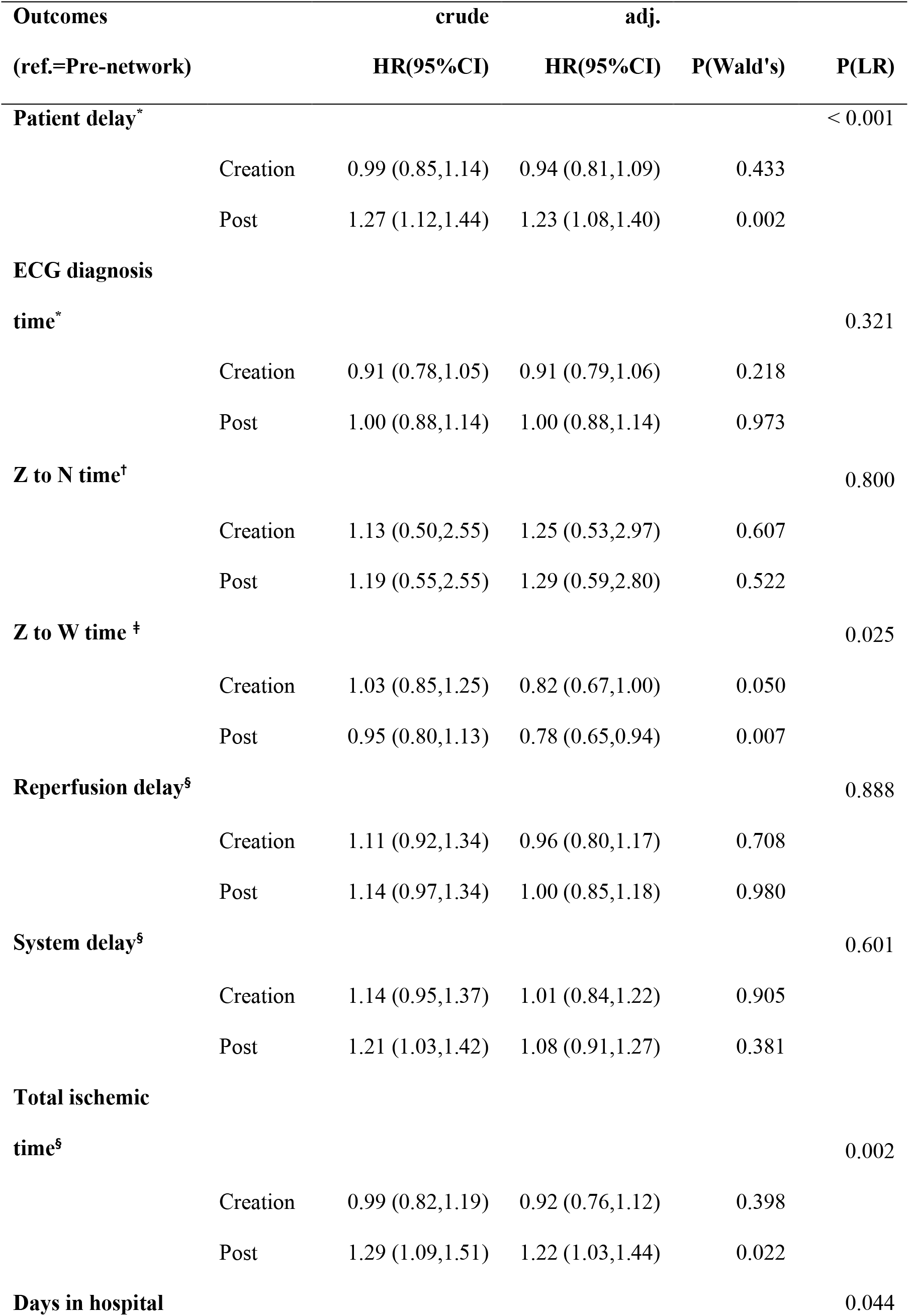
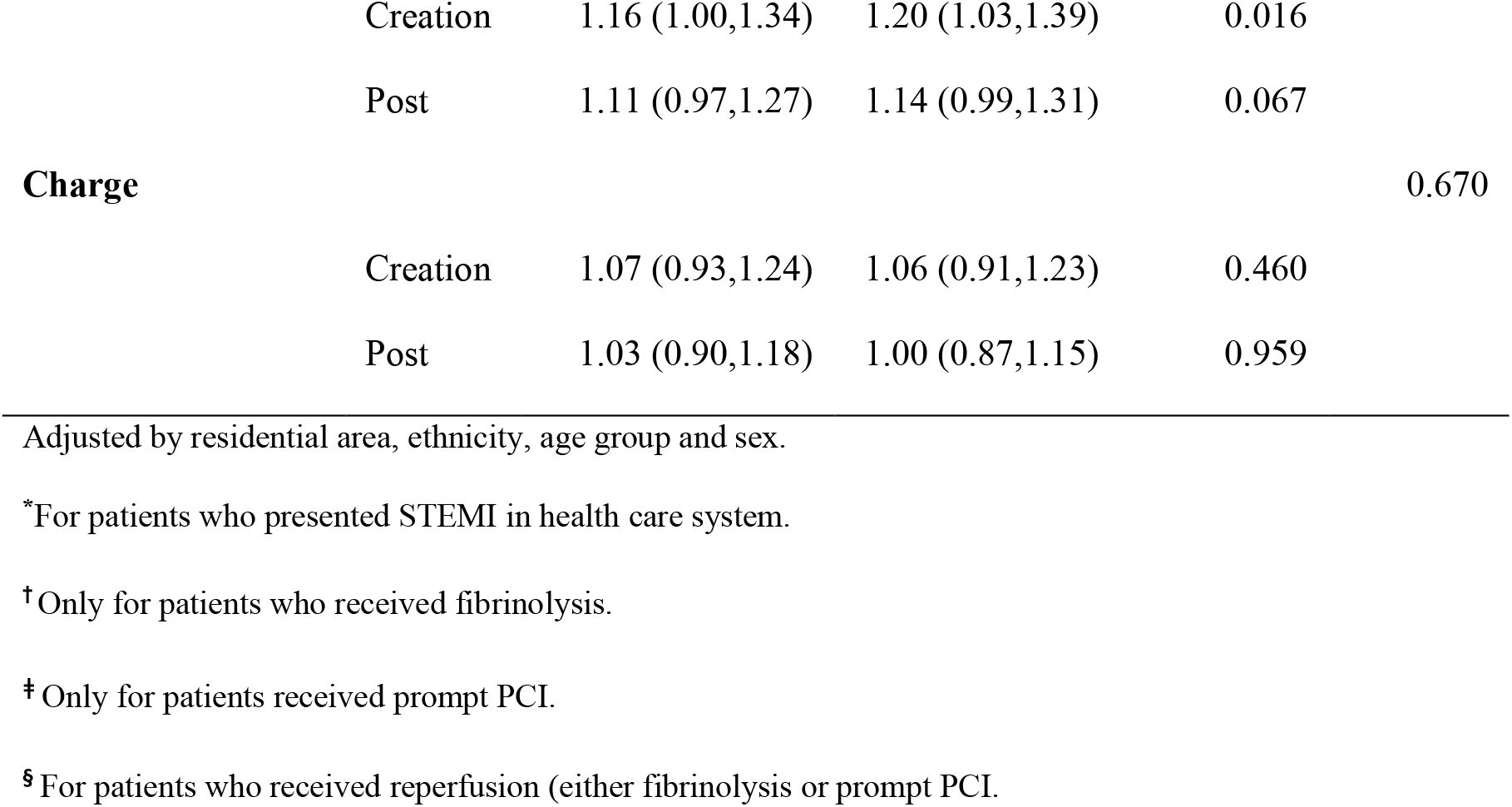
Hazard Ratio of STEMI care outcomes for all patients

Conditioning by area, age, sex and ethnicity, for all patients, compared with pre-network phase, patients during post-network phase were significantly more likely to have shorter patient delay (HR=1.23 [1.08,1.40]); there were nonsignificant HR of diagnosis time for patients during the post-network phase.

For patients received reperfusion (either fibrinolysis or PCI), after conditioning, patients during the post-network phase were significantly more likely to have shorter total ischemic time than the pre-network phase (HR=1.22 [1.03,1.44]); however, there were nonsignificant HR of reperfusion delay for patients during the post-network phase compared with during the pre-network phase.

By conditioning, for patients received fibrinolysis, there was no significant HR of Z to N time between the pre-network patients and the post-network patients. However, for patients received prompt PCI, patients during the post-network phase were significantly less likely to have shorter Z to W time than in the pre-network phase (HR=0.78 [0.65, 0.94).

After conditioning, patients during the creation phase were significantly more likely to have shorter stays in hospital than the pre-network (HR=1.20 [1.03, 1.39]); there was no significant HR of any other continuous outcome variables between the pre-network patients and the post-network patients.

### Competing Risks of Reperfusion Strategies by Network

For fibrinolysis and prompt PCI, once one event occurred for any individual, there is no probability to have another one. Thus, in analysis of time-to-event for each reperfusion method, it was not appropriate to consider patients been censored if they received alternative reperfusion therapy. Instead, competing risks analysis was used to estimate the cumulative incidence of each reperfusion procedure.

The cumulative incidence of PCI and fibrinolysis were shown within 180 minutes as shown in Figure 5, the cumulative incidences of fibrinolysis for all patients were increased by the network, while the cumulative incidence of PCI for all patients were decreased by the network.

**Figure 5.**
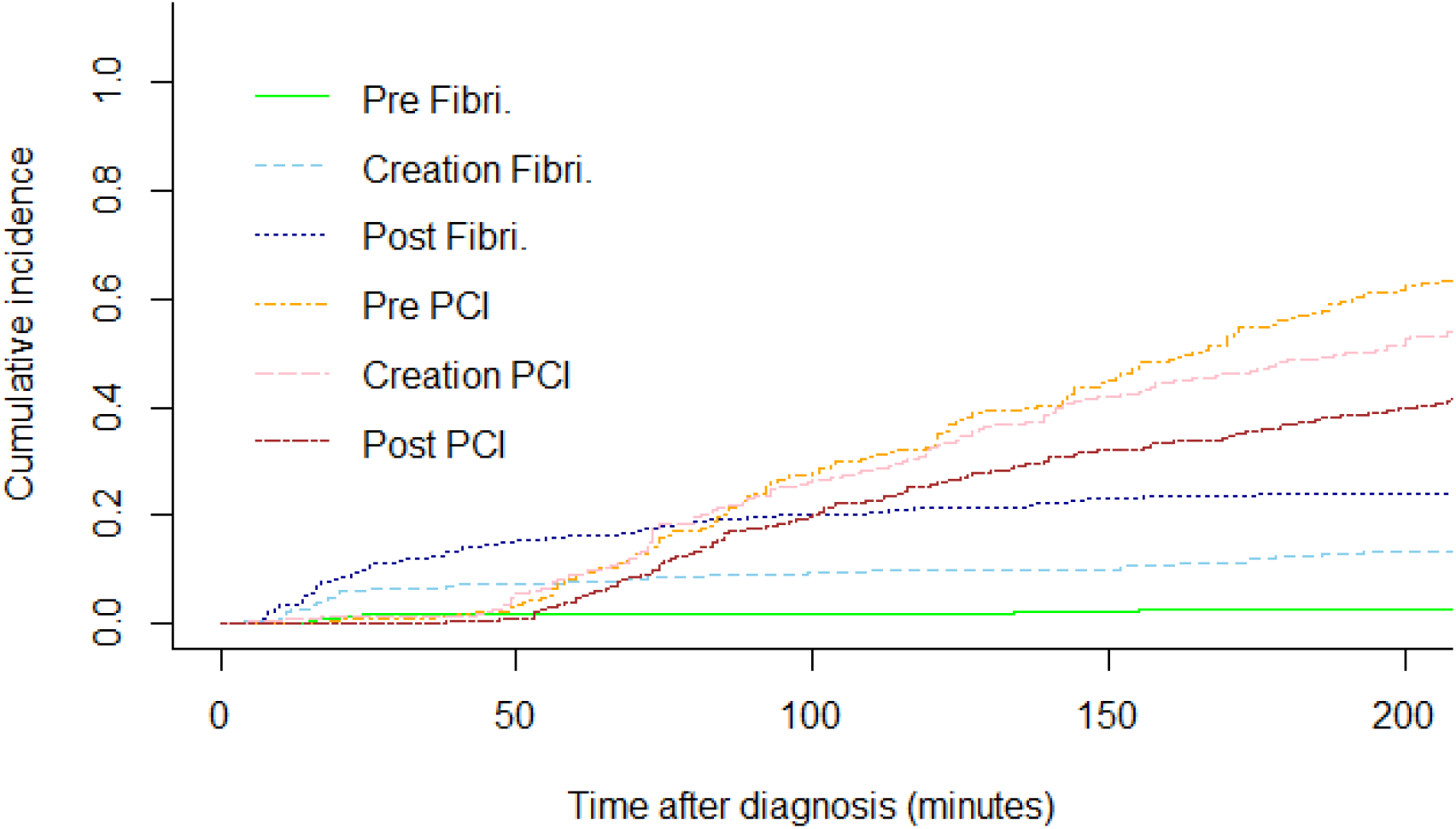
Cumulative incidence of reperfusion strategy by network

The exact cumulative incidences of each procedure at certain timepoints are shown in Table 6. It is noticed that the major reperfusion strategy during the post-network phase was fibrinolysis within 90 minutes after diagnosis but was prompt PCI after 90 minutes, which is consistent with the current guideline for STEMI care.

**Table 6.** Cumulative Incidences of Reperfusion Procedures across Network

| Timepoints (minutes) | Cumulative Incidence |  |  |  |  |  |
| --- | --- | --- | --- | --- | --- | --- |
|  | 30 | 60 | 90 | 120 | 150 | 180 |
| **Fibrinolysis** |  |  |  |  |  |  |
| Pre- | 0.018 | 0.018 | 0.018 | 0.018 | 0.023 | 0.027 |
| Creation | 0.064 | 0.078 | 0.091 | 0.100 | 0.100 | 0.123 |
| Post- | 0.115 | 0.161 | 0.196 | 0.214 | 0.230 | 0.240 |
| **Prompt PCI** |  |  |  |  |  |  |
| Pre- | 0.009 | 0.090 | 0.240 | 0.330 | 0.448 | 0.561 |
| Creation | 0.014 | 0.091 | 0.237 | 0.324 | 0.420 | 0.489 |
| Post- | 0.000 | 0.046 | 0.175 | 0.258 | 0.323 | 0.369 |

## Discussion

### Vulnerable Groups in STEMI Care

Prior the regional Network, rural residents, farmer, female and Yi minority group were evident as the drivers of health inequalities in STEMI care (11). And the atypical patients are usually to be “ignored” either by themselves or by the health care providers. However, following the implementation of the regional network, higher proportions of previously vulnerable groups, especially farmers and atypical patients, were identified by the network.

Compared with the pre-network phase, despite the reduced proportion of rural patients and increased proportion of urban patients, the proportions of farmer markedly increased form 59.7% in pre-network phase to 67.1% in post-network phase, and the proportion of urban farmers during the post-network phase was twice than that in the pre-network phase. Farmer is the major population of China. Even in urban area, there are still many farmers living and working in the sub-urban area. Sub-urban and rural areas usually have much less public resource of community education for STEMI care than central urban area.

One of the major tasks of the regional STEMI network is to promote the public education of STEMI care to increase health awareness in community, especially focusing on sub-urban and rural areas. The public education of the network also focusses on the introduction of the atypical symptom in the community, and the professional training to identify atypical symptom for health care providers. Live speeches, promotion videos, brochure pamphlets and leaflets were employed by the professional training teams around almost all the city area and each county to propagandize the public STEMI care knowledge. Everyone could participate in the speech and take away the brochures for free. In almost all remote rural area, the related poster with more picture than words were posted on the wall in the central area of the villages.

The effectiveness of public education by the network was evident in the current study. The influence of regional STEMI network on farmers’ awareness might be the reason for the increased proportion of farmers. On the other hand, by the professional training of the regional STEMI network, the STEMI care ability in hospitals were improved, especially for doctors in county and town hospitals. Higher proportion of farmers and patients with atypical symptom were identified following the Network implementation. Although there is no other study focusing on it, it is really a pleasing finding for the clinical field. The more atypical patients are identified, the more health inequality could be discovered and eliminated.

### Timely presentation and shorter patient delay

Presentation mostly depends on the patients, care givers and the accessibility of the local health service (such as ECG in health care unit). However, the awareness of seeking medical service during the pre-network phase was limited. Many people living sub-urban and rural area did not know they should seek emergency medical service when they suffered from serious chest pain. And, before the implementation of the regional Network, many health personnel who work in a town hospital or primary care unit do not have the ability to immediately recognize and diagnose STEMI.

“At once Chest Pain, Seeking EMS Immediately”, which is printed in the brochure pamphlet and leaflet, is the theme of the public education of the regional STEMI network. On the other hand, enhancing the ability of diagnosing STEMI by ECG and myocardial enzyme is one of the major tasks of the Network. Following the implementation of the regional STEMI network, professional training programs such as “Identifying STEMI by ECG” were conducted in each county and/or town hospital. All county hospitals and more than half of the town hospitals had set up the laboratory for “Rapid Detection of Myocardial Enzyme”. For some health care providers who cannot recognize ECG precisely, consultation with experts’ team from Network central hospital were provided online or by smartphone with 24 hours service. Hence, all health care units in villages, towns and counties had the ability to diagnose STEMI in 10 minutes. For those patients without ST-segment change in first 10 minutes after FMC, more frequently ECG monitoring were conducted during the post-network phase. Hence, there was a higher probability to identify STEMI early than before. The efficiency of public education by the regional Network on timely presentation was evident in the current study.

A study reported the eastern Austrian STEMI network (12), which serves a population of approximate 766 000 inhabitants within a region of 4186 km^2^, and comprises 20 pre-hospital emergency medical service units, 4 hospitals and 3 cardiac intervention centres. Treatment guidelines were updated in 2012 and documentation within a web-based STEMI registry became mandatory. A total of 416 STEMI cases from February 2012 to April 2015 were documented, the median time loss between onset of pain and EMS call was 54 minutes, while pre-hospital delay was 46 minutes.

### Higher reperfusion rate and shorter ischemic time

After first diagnosis of STEMI, the quality of STEMI reperfusion care mainly depends on the specific reperfusion care accessibility of health care unit and health care providers. Before the implementation of the Network, both prompt PCI and fibrinolysis are not available in county/town hospitals and health care units. Suspected STEMI patients might be transported from a primary care unit to a town hospital, and subsequently be transported from the town hospital to a county hospital, finally arriving at the prefecture-level hospital, which can provide primary PCI. Precious time is wasted due to “step-by-step transportation” from non-PCI hospitals to catheter room.

However, promoting fibrinolysis as reperfusion priority strategy in county/town hospitals and health care units is one of the major tasks of the regional Network. By popularization the fibrinolysis, more health care providers in county/town hospitals and health care units could provide fibrinolysis as emergency reperfusion to patients, which markedly increased the total proportion of reperfusion patients.

After the implementation of the Network, there was a higher proportion of reperfusion (65.3%) than during the pre-network phase (58.2%) with an OR 1.33. The finding is consistent with another study (13) assessed the effectiveness of the regional Network on STEMI care in a developed area of China. The study included 165 patients before and 343 cases at 1 year after implementation of the network. Compared to before, the reperfusion rate was significantly increased after establishment of the network (84.5% vs 75.5%, P = 0.06). Several studies from western countries also reported similar findings. One study from Romania (14), analysed 5,899 consecutive patients with STEMI between 2004 and 2011; introduction of the network was associated with an absolute change in the use of reperfusion therapy (87.15% vs. 26.94%, P < 0.001) for patients presenting within 12 hours after the onset of symptoms. Another study in Poland (15) reported a 24-hour primary PCI regional service, and showed that reperfusion therapy was used in 48.7% of STEMI patients before regional service, but in all patients after the regional service.

### Improvement of fibrinolysis

In the study location, there is only one tertiary hospital that can provide 24-hours prompt PCI. According to the guideline, emergency fibrinolysis should be provided to patients if the PCI is not available in time. However, before the implementation of the Network, fibrinolysis was unavailable in all town hospitals and primary care units in under-developed areas. The reasons for low fibrinolysis rate maybe include: the patients knew little about fibrinolysis, and/or worried about the risk of fibrinolysis, and the local health unit lacked fibrinolysis agent and/or the ability of medical personnel to perform emergency reperfusion care, etc.

The major task of the regional network is to popularize the fibrinolysis as priority reperfusion strategy in non-PCI hospital. Professional training programs such as “Benefits and Risks of fibrinolysis, Monitoring Complications” were conducted by the training team in each county and/or town hospital, examinations of the performance of fibrinolysis were conducted by the headquarters of the regional network in each non-PCI hospital, regularly and/or randomly. By the implementation of the regional network, Fibrinolysis was efficiently conducted in remote rural areas. More than half of the town hospitals had set up the laboratory for “Rapid Detection of Myocardial Enzyme” and a storeroom for fibrinolytic agents. Increasing proportions of fibrinolysis were discovered from the pre-network phase to the post-network phase. The competing risks analysis also showed an increasing trend in cumulative incidence of fibrinolysis following the Network implementation. Especially within the first 90 minutes after diagnosis, fibrinolysis already was the major strategy for STEMI patients during the post-network phase.

“Late but better than None”, even there was only a little achievement of timely fibrinolysis, increasing the fibrinolysis rate is always the first step to improve reperfusion therapy in non-PCI hospital. Though less than one quarter patients received emergency fibrinolysis therapy, the median Z to N time in current study were much shorter with a median of 38 [17-86] minutes. A Scotland study (16) in 2018 reported that the median time from first medical contact to initiation of fibrinolysis for rural patients was 125 (range 104 to 140) minutes while for patients from urban area was 80 (range 78 to 93) minutes.

### Delayed Reperfusion

Despite the total ischemic time of patients during the post-network being significantly shorter and the reperfusion rate higher than during the pre-network phase, the reperfusion delay was not significantly improved. Compared with the patients during the pre-network phase, there were lower proportions of timely PCI and timely reperfusion for patients during the post-network.

Timely reperfusions comprise timely PCI and timely fibrinolysis. Timely PCI is defined as Z to W time less than 90 minutes and timely fibrinolysis is defined as Z to N time less than 10 minutes in the current study according to the ESC guideline (17). It is really an extremely minimal time for the real clinical work, especially in under-developed area. In the modified protocol for STEMI care from China Chest Pain Centre (18), timely PCI is defined as the time between FMC and PCI wire-crossing of less than 120 minutes; timely fibrinolysis is defined as the time between FMC and fibrinolysis needle time of less than 30 minutes. The modified protocol is more appropriate for developing and under-developed area.

Though emergency fibrinolysis was promoted in rural area by the regional STEMI network, prompt PCI is still the major reperfusion strategy for all patients. During the post-network phase, there were around 47% patients received PCI while only 18% cases received fibrinolysis. According to the analysis of the background of patients, a higher proportion of urban farmer was found during the post-network phase (22%) than the pre-network (9%). Urban farmers usually live in sub-urban area. The fact that 90% of Chuxiong prefecture is mountainous hinders the situation. The major challenge is that driving times from most sub-urban and rural areas to the only PCI centre are usually more than 90-120 minutes, which is the maximal allowable time for transferring patients to receive primary PCI. Hence, if more urban farmers had first medical contact and received the diagnosis of STEMI in rural area, and most of them preferred to receive prompt PCI as their reperfusion therapy, it is unavoidable to have longer Z to W time.

Setting up PCI catheter room in local area is the efficient way to shorten Z to W time; however, it is not a reality in an under-developed area in the long term. Huge financial support and professional cardiologist are necessary to enhance the accessibility of PCI in suburban areas. Before that, based on the current situation in rural and mountainous areas, “bidirectional transportation” and “mobile fibrinolysis during transportation” are the priority for the regional STEMI network in Chuxiong prefecture.

### Limitations and Strengths

One of the limitations of the study is that a few STEMI patients might be outside the registered system. And patients might die before medical contact (at home or on the way to hospital). Only the STEMI patients who had medical contact were included in the current study.

Nevertheless, strengths of current study should be noted. The current study is a 5-year prefecture-wide study covering the whole phase of setting up the regional STEMI network; it is well representative of patients who have medical contact. All the data are from the real world setting without any human disturbance. The data source into Chest Pain Centre of Chuxiong prefecture has already been certified by the headquarters of China Chest Pain Centre, which is authentic and credible. Furthermore, it is the first description of STEMI patients undergoing STEMI network implementation with large sample size in an under-developed area.

## Conclusion

Among STEMI patients who had medical contact, there were higher proportion of farmer and atypical patients during the post-network phase than during the pre-network network.

Following the implementation of the regional network, among all STEMI patients, higher proportions of timely presentation, reperfusion and shorter patient delay were found during the post-network phase than during the pre-network phase. After conditioning, patients during the post-network phase were more likely to have timely presentation, reperfusion therapy and shorter patient delay. There were no significant difference of heart failure or mortality among the three phases.

Among patients who received reperfusion (either prompt PCI or fibrinolysis), shorter system delay and total ischemic time were found for post-network patients than pre-network patients. However, after conditioning, for patients who received reperfusion, those during post-network phases were less likely to receive timely reperfusion than patients during the pre-network phase; for patients who received prompt PCI, the patients during post-network phases were less likely to receive timely PCI and less likely to have shorter Z to W time than patients during the pre-network phase.

The cumulative incidence was increased for fibrinolysis while decreased for prompt PCI by the network. The major reperfusion strategy during the post-network phase was fibrinolysis within 90 minutes after diagnosis but prompt PCI after 90 minutes.

Improvements of STEMI reperfusion care by the regional Network were evident in this under-developed area; however, timely reperfusion care still needs to be enhanced.

## Data Availability

Will individual participant data be available (including data dictionaries)? Yes. What data in particular will be shared? Individual participant data that underlie the results reported in this article, after de-identification (text, tables, figures, and appendices). What other documents will be available? Study Protocol, Statistical Analysis Plan, Informed Consent Form, Clinical Study Report, Analytic Code. When will data be available (start and end dates)? Beginning 6 months and ending 36 months following article publication. With whom? Investigators whose proposed use of the data has been approved by an independent review committee identified for this purpose. For what types of analyses? To achieve aims in the approved proposal, such as to compare with STEMI care in other region. By what mechanism will data be made available? Proposals should be directed to the responsible author. To gain access, data requestors will need to sign a data access agreement.

## Acknowledgements

We thank all the staff of the Chest Pain Center Alliance of Chuxiong prefecture, Yunnan, China, for their hard work and selfless contributions to STEMI care.

## Source Funding

None.

## Disclosures

All authors declare that they have no competing interests.

## Notes

**Source(s) of support/funding:** People’s Hospital of Chuxiong Prefecture, Yunnan, China.

### Competing Interest Statement

The authors have declared no competing interest.

### Clinical Trial

It is a retrospective observational study.

### Author Declarations

After receiving the ethical approval from the Chest Pain Centre of Chuxiong prefecture, Yunnan, China, and also from the Human Research Ethic Committee in the Prince of Songkhla University, Thailand, the STEMI registry system database was searched to access STEMI index cases.

## References

1. Yeh RW, Sidney S, Chandra M, Sorel M, Selby JV, Go AS. Population trends in the incidence and outcomes of acute myocardial infarction. N Engl J Med. 2010 Jun 10;362(23):2155–65.

2. Gupta R, Joshi P, Mohan V, Reddy KS, Yusuf S. Epidemiology and causation of coronary heart disease and stroke in India. Heart Br Card Soc. 2008 Jan;94(1):16– 26.

3. Zhang X, Khan AA, Haq EU, Rahim A, Hu D, Attia J, et al. Increasing mortality from ischaemic heart disease in China from 2004 to 2010: disproportionate rise in rural areas and elderly subjects. 438 million person-years follow-up. Eur Heart J Qual Care Clin Outcomes. 2017 Jan 1;3(1):47–52.

4. Li J, Li X, Wang Q, Hu S, Wang Y, Masoudi FA, et al. ST-segment elevation myocardial infarction in China from 2001 to 2011 (the China PEACE-Retrospective Acute Myocardial Infarction Study): a retrospective analysis of hospital data. Lancet. 2015 Jan 31;385(9966):441–51.

5. Ibánez B, James S, Agewall S, Antunes MJ, Bucciarelli-Ducci C, Bueno H, et al. 2017 ESC Guidelines for the management of acute myocardial infarction in patients presenting with ST-segment elevation. Rev Espanola Cardiol Engl Ed. 2017 Dec;70(12):1082.

6. Benedek I, Gyongyosi M, Benedek T. A prospective regional registry of ST-elevation myocardial infarction in Central Romania: Impact of the Stent for Life Initiative recommendations on patient outcomes. Am Heart J. 2013 Sep 1;166(3):457–65.

7. Januś B, Rakowski T, Dziewierz A, Fijorek K, Sokołowski A, Dudek D. Effect of introducing a regional 24/7 primary percutaneous coronary intervention service network on treatment outcomes in patients with ST segment elevation myocardial infarction. Kardiol Pol. 2015;73(5):323–30.

8. Wang B, Wang Y, Ye T, Xiao G, Chang H, Wen H, et al. [Integrated regional network construction for ST-segment elevation myocardial infarction care]. Zhonghua Xin Xue Guan Bing Za Zhi. 2014 Aug;42(8):650–4.

9. http://www.cxz.gov.cn/info/egovinfo/1001/xxgkxt_content/115323000151673132-/2019-0429001.htm. In.

10. https://finance.sina.com.cn/roll/2019-01-15/doc-ihqfskcn7140929.shtml. In.

11. Zhang LM, Geater AF, McNeil EB, Lin YP, Liu SC, Luo H, et al. Health Inequalities of STEMI Care Before Implementation of a New Regional Network: A Prefecture-Level Analysis of Social Determinants of Healthcare in Yunnan, China. Int J Health Policy Manag [Internet]. 2021 May 11 [cited 2021 Jun 25];0. Available from: https://www.ijhpm.com/article_4047.html

12. Emergency management of patients with ST-segment elevation myocardial infarction in Eastern Austria: a descriptive quality control study. - PubMed - NCBI [Internet]. [cited 2019 Jun 12]. Available from: https://www.ncbi.nlm.nih.gov/pubmed/?term=Emergency+management+of+patients+with+ST-segment+elevation+myocardial+infarction+in+Eastern+Austria+3A+a+descriptive+quality+control+study

13. Wang B, Wang Y, Ye T, Xiao G, Chang H, Wen H, et al. [Integrated regional network construction for ST-segment elevation myocardial infarction care]. Zhonghua Xin Xue Guan Bing Za Zhi. 2014 Aug;42(8):650–4.

14. Benedek I, Gyongyosi M, Benedek T. A prospective regional registry of ST-elevation myocardial infarction in Central Romania: impact of the Stent for Life Initiative recommendations on patient outcomes. Am Heart J. 2013 Sep;166(3):457–65.

15. Effect of introducing a regional 24/7 primary percutaneous coronary intervention service network on treatment outcomes in patients with ST segment … - PubMed - NCBI [Internet]. [cited 2019 Jun 12].

16. Pedley DK, Bissett K, Connolly EM, Goodman CG, Golding I, Pringle TH, et al. Prospective observational cohort study of time saved by prehospital thrombolysis for ST elevation myocardial infarction delivered by paramedics. BMJ. 2003 Jul 5;327(7405):22–6.

17. ESC Guidelines for the management of acute myocardial infarction in patients presenting with ST-segment elevation | European Heart Journal | Oxford Academic [Internet]. [cited 2021 May 15]. Available from: https://academic.oup.com/eurheartj/article/33/20/2569/447818

18. Xiang D, Xiang X, Zhang W, Yi S, Zhang J, Gu X, et al. Management and Outcomes of Patients with STEMI During the COVID-19 Pandemic in China. J Am Coll Cardiol. 2020 Sep 15;76(11):1318–24.

